# Investigation of in-phase bilateral exercise effects on corticospinal plasticity in relapsing remitting multiple sclerosis: a multiple baseline design

**DOI:** 10.1101/2022.07.14.22277636

**Authors:** Dimitris Sokratous, Charalambos C. Charalambous, Eleni Zamba-Papanicolaou, Kyriaki Michailidou, Nikos Konstantinou

**Author notes:** **Corresponding Author:** (DS). **Authors’ contributions:** D.S., C.C.C., E.Z.P., and N.K., are responsible for the conception and the experimental design. D.S., and K.M., are responsible to collect, analyse and interpret the data. D.S., and N.K., are responsible to draft the manuscript. N.K., C.C.C., and K.M., revised the manuscript critically for important intellectual content. ClinicalTrials.gov NCT05367947.

## Abstract

Relapsing-remitting multiple sclerosis is the most common type of multiple sclerosis characterized by periods of relapses and generating various motor symptoms. These symptoms are associated with the corticospinal tract integrity, which is quantified by means of corticospinal plasticity which can be probed via transcranial magnetic stimulation and assessed with motor threshold, motor evoked potential and central motor conduction time. Several factors, such as exercise and interlimb coordination, can influence corticospinal plasticity. Previous work in healthy and in stroke patients showed that the greatest improvement in corticospinal plasticity occurred during in-phase bilateral arm exercises. Altered corticospinal plasticity due to bilateral cortical lesions is common in multiple sclerosis, yet, the impact of these type of exercises in this cohort is unclear. The aim of this concurrent multiple baseline design study is to investigate the effects of in-phase bilateral exercises on corticospinal plasticity and on clinical measures using transcranial magnetic stimulation and standardized clinical assessment, in five people with relapsing-remitting multiple sclerosis. The intervention protocol will last for 12 consecutive weeks (30-60 minutes /session x 3 sessions/week) and include in-phase bilateral movements of the upper limbs, adapted to different sports activities and to functional training. To define functional relation between the intervention and the results on corticospinal plasticity (i.e., resting motor threshold, motor evoked potential amplitude, latency) and on clinical measures (i.e., balance, gait, bilateral hand dexterity and strength, cognitive function), we will perform a visual analysis followed by multilevel modelling and the single case educational design-specific mean difference in order to estimate the magnitude of the effect size across cases. We assume that possible effects from our study, will introduce a type of exercise that will be effective during the disease progression.

## Introduction

Multiple sclerosis (MS) is the most common inflammatory demyelinating and neurodegenerative disease of the central nervous system (1). The global prevalence of MS during the last decade has increased by 30%, while the number of people suffering with MS worldwide is estimated at approximately 2.8 million(8). The low mean age of diagnosis (i.e., 32 years old), along with an average of seven years’ shorter life expectancy (i.e., 74.7 years) compared to the general population (4–6), highlights the need for a lengthy support, resulting in increased financial burden (7). The direct cost from health care services and the indirect cost due to productivity loss because of physical inactivity of people with MS, indicates a huge financial burden on families, healthcare systems, and societies in general. Recent studies reported that the annual mean cost of health care systems for people with MS living in Europe is about €40,000 (8). Additionally, both MS patients and their caregivers, who usually are family members, are facing several psychological and social difficulties due to social isolation, poorer quality of life, reduced productivity and lower general health levels (9,10).

Relapsing-remitting multiple sclerosis (RRMS), is the most common type of MS and is characterised by periods of relapses followed by partial or complete recovery (11). Inflammatory lesions are commonly found bilaterally in both white and grey matter of the central nervous system (12,13), resulting in diverse clinical condition and symptoms, that include motor and cognitive impairments, vision deficits, depression and fatigue (13–15). Those symptoms results in significantly low quality of life (16,17), which subsequently causes the need for a lifelong support and management of symptoms for most people with RRMS (18).

Motor symptoms in RRMS are associated with changes in corticospinal tract integrity and neuroplasticity (19–25). The corticospinal tract is one of the major motor descending pathways providing voluntary motor function in humans (26). The neuroplasticity of the corticospinal tract, as defined by changes in neuron structure or function detected either directly from measures of individual neurons or inferred from measures taken across populations of neurons (27), is an essential factor that predicts clinical recovery in the post-relapse stage of people with RRMS (28,29). Corticospinal plasticity can be probed using Transcranial Magnetic Stimulation (TMS) (30,31) and characterized via certain TMS-specific neurophysiological measures including resting motor threshold, motor-evoked potential (MEP) amplitudes and the central motor conduction time (31). Motor threshold and MEP are the hallmark measures of corticospinal excitability in MS (32), whereas the central motor conduction time expresses the time taken for neural impulses to travel over the central nervous system on their way to the target muscle (33).

Corticospinal plasticity is exercise-dependent (34,35) and influenced by various factors (36,37), such as aerobic exercise (22,38–40), resistance training (22,40), as well as interlimb coordination (41,42). Previous studies that assessed corticospinal plasticity using TMS in healthy participants and in chronic stroke survivors, reported that interlimb coordination and especially in-phase bilateral movement has the strongest effect on corticospinal plasticity (43–46). These effects are thought to be due to the suppression of cortical inhibition (46,47) and the simultaneous activation of homologous representations of the motor cortices, which involves interhemispheric facilitation via transcallosal connection between the primary motor cortex and the supplementary motor area (48,49).

Despite the broad literature on the effects of different types of exercises on the neuroplasticity in people with RRMS (39,50–52), it is unclear whether in-phase bilateral exercises can promote motor related neuroplastic changes in RRMS. In light of evidence that people with RRMS have bilateral cortical lesions (53) which cause bilateral changes of corticospinal tract integrity (23,25), these findings raise the question about the effects of in-phase bilateral exercises on corticospinal plasticity. Such effects would provide strong evidence about whether exercise, in particular in-phase bilateral exercise, can influence the corticospinal plasticity in RRMS.

The aim of this study is to investigate whether a 12-week intervention protocol of in-phase bilateral exercises (independent variable) for the upper limbs, which are adapted to sports activities and to functional training, can significantly affect the corticospinal plasticity and subsequently the individual clinical condition of people with RRMS. Our primary hypothesis is that a significant improvement of corticospinal plasticity will be detected bilaterally, caused by the specific intervention protocol which includes in-phase bilateral exercises of the upper limbs in people with RRMS. We will assess the corticospinal plasticity bilaterally using TMS and calculate TMS-specific measures (45). Visual analysis will be conducted separately for each variable and results will be presented graphically according to the level, trend, and stability, to define functional relationships between the intervention protocol and the corticospinal plasticity. Also, quantitative analysis within cases will be conducted using Cohen’s d, Hedges’ g and piecewise regression analysis, as well as quantitative analysis will be performed between cases, using a multilevel modelling. To estimate the overall effect size, we will use the single case educational design, and the mean difference index (54).

Exploratory analyses in Stage 2 will investigate the effects of the specific exercises protocol on clinical symptoms using clinical assessment (i.e., gait, balance, strength, hand dexterity, cognitive function) and the Modified Fatigue Impact Scale (55). Visual and quantitative analysis will be contacted as described above.

The study follows a concurrent multiple baseline design across subjects (56), which involves five people with RRMS. The specific design has the advantage to verify the cause-effect inference clearly by the staggered duration through separate baseline phases (57). Consequently, we assume that possible effects from our study will promote a novel approach in the field of neurorehabilitation and will introduce a type of exercise that will be effective during the disease progression.

## Materials and Methods

### Participants

All participants will be recruited from August to October 2022 and evaluated by an experienced neurologist at The Cyprus Institute of Neurology and Genetics. The inclusion criteria include 1) diagnosed with RRMS, 2) Expanded Disability Status Scale score between three and five (58), 3) aged between 30 and 70 years, 4) no relapse within 30 days and 5) Mini Mental State of Examination score between 24 and 30 (no cognitive impairment) (59). The exclusion criteria include 1) metal implants, 2) history of any disease affecting the central nervous system other than MS, 3) history of cardiovascular disease, 4) mental disorders, and 5) severe orthopaedic disorders, 6) pregnancy, 7) visual deficit, 8) hearing impairments, 9) epileptic seizures and 10) spasticity level on upper or lower limbs more than 1+ (slight increase in muscle tone) according to Modified Ashworth Scale (60). Additionally, participants will be advised to continue their usual prescribed medication throughout the study duration, and they will be advised to continue their usual routine avoiding receiving any other exercise program during this study. Furthermore, all participants will read and sign a written informed consent while all procedures are approved and conducted in accordance with the ethical guidelines of the Cyprus National Bioethics Committee before recruitment.

### Study design

The study follows a concurrent multiple baseline design across subjects, without blinding and has been designed according to the ‘What Works Clearinghouse’ criteria for single case studies (56). According to Kratochwill et al. (56), three participants, with collection of three data points across different phases is the minimum number needed to meet the standard criteria, while four or more is recognized as more reliable. We aim to include five participants to ensure the reliability of the results in case of dropouts, as well as to record several data points across the baseline phase, five data points during the intervention phase and three data points in the follow up phase. During the experimental procedure, all participants will begin the study with the baseline phase at the same time while the intervention phase is introduced staggered across patients and time (Fig 1). The intervention will be introduced systematically in one patient while baseline data collection continues in the others without any intervention. The cause-effect inference can be clearly verified by the staggered duration through separate baseline phases (57). Subsequently, if the intervention (i.e., in-phase bilateral exercises) is the sole cause of improvement in participants’ conditions, the proposed outcome measures will not change for the participants that remain in the baseline phase but will be improved only for those in the intervention phase.

**Figure 1.**
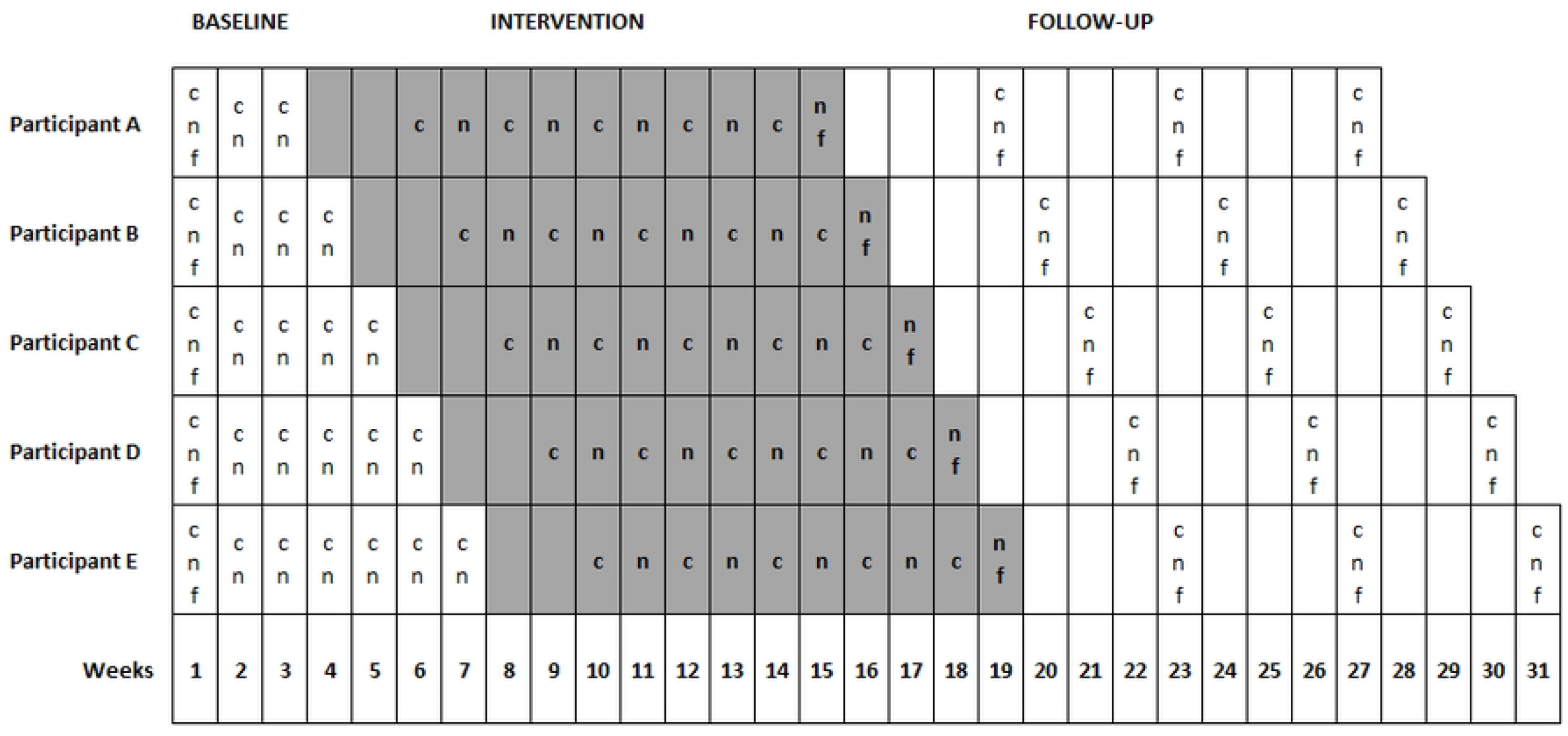
Timeline and schematic representation of the study’s design. Grey colour represents the intervention phase. Each row (A-E) represents a different participant. (c) clinical assessment. (n) neurophysiological assessment via TMS. (f) Modified Fatigue Impact Scale questionnaire. Every cell represents a different week, so every procedure which is included (i.e., c, n, f) will be performed during the corresponding week but in different days.

### Baseline

As depicted in Figure 1, all patients will begin the baseline phase simultaneously. Each patient will undergo a baseline phase of a different time duration, starting with three weeks for the first participant and gradually increasing by one week for each participant. During the baseline phase, each participant will be assessed on the Modified Fatigue Impact Scale (55) on the first week. The neurophysiological (primary outcome measures) and clinical (secondary outcome measures) assessments will be repeated each baseline week for all participants.

### Intervention

Immediately after the end of each baseline phase, the intervention phase will begin staggered across participants and time accordingly (Fig 1). The intervention protocol consists of exercises based on in-phase bilateral movements of the upper limbs, which are adapted to different sport activities and to fitness functional exercises, organized in a circuit training considering the MS exercise recommendations (61). Since no established protocols have been previously reported, for the needs of our study a certified fitness instructor will design these protocols adapted to different sport activities. Specifically, each session will consist of one to three sets, consisting of 10–15 repetitions of 9 different exercises targeting large muscle groups of the upper limbs (shoulder flexors, extensors, rotators, abductors and adductors, elbow flexors and extensors, hand and finger flexors and extensors). Additionally, three exercises will target large lower limb muscle groups (hip flexors, extensors, abductors and adductors, knee and ankle flexors and extensors) to be performed in between the upper limbs exercises to allow relaxation of the upper limb muscles.

The specific exercises will include sports activities of basic technical skills of basketball (e.g., different types of passing, catching and throwing the ball) and volleyball (e.g., different types of passing and receiving the ball), whereas the fitness exercises will include shoulder rows, shoulder lateral raises, elbow flexions, elbow extensions, as well as using the diagonal movements form proprioceptive neuromuscular facilitation technique (62) by the use of resistance elastic bands (51), as well as exercises with the patients’ own body weight (e.g., pushups, TRX) (63). To maintain the interest of the participants, the exercise program will be modified throughout the course of the 12-week intervention period via changing the level of difficulty. For example, elastic bands with different resistance levels will be used, the number of repetitions and sets will vary along with the specific exercise and body position (e.g., from sitting to standing).

The intervention phase for each participant will consist of 12 consecutive weeks in which the proposed protocol will be performed three times per week, for 30-60 minutes each session, adapted to each participant’s fatigue and fitness level. Each participant has to complete at least 27 (75%) out of 36 sessions in order to be included in the data analysis (51). Every intervention session will consist of a five minutes’ warm-up (i.e., whole body range of motion exercises), followed by the main sport activities and fitness exercise protocol as described above, and a cool down session for five minutes (i.e., passive stretching exercises of the muscle groups which are involved in the main part).

Additionally, starting from the third intervention week, we will perform five neurophysiological (see primary outcome measures) and five clinical assessments (see secondary outcome measures) (i.e., once a week), to collect five data points for every participant across the intervention phase. Moreover, each participant will also be asked to complete once the Modified Fatigue Impact Scale (see secondary measures) at the end of the intervention phase (51) (Fig 1).

### Follow-Up

As depicted in Figure 1, every participant will undergo three follow-up assessments in total, after finishing the training protocol lasting from February to May 2022, so to explore possible long-lasting effects. Each follow-up assessment includes both neurophysiological and clinical measures (including Modified Impact Scale). We will perform the first follow-up assessment at the end of the fourth post-intervention week, the second one at the end of the eighth post-intervention week and we will perform the last follow-up assessment at the end of the 12^th^ post-intervention week (Fig 1).

## Data Acquisition of Outcome Measures

### Primary Outcome Measures

We will assess the corticospinal plasticity using single pulse TMS in the neurophysiology lab of the Cyprus Institute of Neurology and Genetics. Using electromyography (EMG) signals, we will analyse bilateral cortical excitability and bilateral central motor conduction time to determine corticospinal plasticity and therefore to test the primary hypothesis. The resting motor threshold and the MEP amplitude of Abductor Pollicis Brevis muscle will define cortical excitability, while we will use the MEP latency to calculate the central motor conduction time (see below). During all neurophysiological assessments, participants will be in a relaxed sitting position in a comfortable chair with feet touching the floor and both arms will be placed on cushioned armrests and with the head rested on a cushion. To ensure methodological consistency, we will collect all data by performing the same methodological procedures for both conditions (i.e., cortical excitability and central motor conduction time) bilaterally (one side per assessment), across participants and across all time points.

### EMG recording

During both TMS and peripheral stimulation, surface EMG of the Abductor Pollicis Brevis muscle will be collected. We will follow a standard skin preparation (64) and surface disk electrodes placement procedures by attaching the electrodes over the end plate region of the Abductor Pollicis Brevis muscle (65). Specifically, the anode (red colour) electrode will be placed distally, whereas the cathode (black colour) electrode proximally. A ground reference electrode will be attached on the lateral condyle of the elbow, of the corresponding upper limb. Additionally, all signals will be filtered with the help of the KeyPoint Net Software Electromyography using a bandwidth of 2 Hz–10 kHz, 1mV/D for the MEP sensitivity with a single-pulse stimulation frequency.

### Peripheral stimulation

In addition to MEP latency, calculating the central motor conduction time requires two peripheral derived measures, the F wave (i.e., late muscle response) and the M wave (i.e., direct muscle response) (66,67). Therefore, we will initially deliver peripheral stimulation on the median nerve at the wrist, approximately in 8 cm distance from the cathode electrode (65), while collecting EMG from the Abductor Pollicis Brevis muscle (68) of the corresponding upper limb. In order to determine possible changes in central motor conduction time, we will analyse F and M wave (69).

### TMS assessment

Following TMS recommended guidelines concerning safety and experimental conditions (70,71), we will assess bilateral cortical excitability and bilateral central motor conduction time. We will apply TMS single pulses (72) via figure-eight coil (C-B60; inner diameter: 35mm, outer diameter: 75mm), connected to the MagPro R20 (MagVenture User Guide, United Kingdom edition, MagVenture A/S, Denmark). The coil will be oriented tangentially over the contralateral motor area of the brain, relative to the target muscle (i.e., Abductor Pollicis Brevis), with a posterolateral handle pointing in approximately 45 degrees angle to the sagittal plane, as a result to induce current in a posterior-anterior direction in the brain (73).

For the TMS procedures, we will first find the optimal stimulation site (i.e., hot-spot), next we will determine the resting motor threshold, and then apply a bout of single pulses using suprathreshold stimulation. To determine hot-spot (i.e., the spot in which the largest response of the target muscle is elicited), we will deliver single pulses at low intensities (e.g., ∼20% maximum stimulator output) and gradually increase it by 5%, maximum stimulator output until we will reach the intensity that will elicit three consecutive MEPs with peak-to-peak amplitude greater than 50mV (74,75). Then, we will mark the position of the coil on the skull with a water-resistant ink, so to determine the resting motor threshold of the target muscle. Resting motor threshold is the minimum stimulation intensity needed to produce MEPs of the target muscle, which is defined by the hot-spot. To identify the resting motor threshold of the Abductor Pollicis Brevis muscle, we will employ an adaptive threshold-hunting method, the Motor Threshold Assessment Tool (MTAT 2.0) (76) (available at http://clinicalresearcher.org/software.htm). The specific method has the advantage of speed without losing accuracy when compared to the relative-frequency methods based on the Rossini–Rothwell, although both methods have similar precision (77). Then, to quantify the MEP-derived measures of interest (i.e., MEP amplitude, latency), we will apply 25 suprathreshold stimuli (78) at 120% of the resting motor threshold (79).

### Secondary Outcome Measures

The secondary outcome measures will include all the clinical assessments, which will be performed in the physiotherapy unit of the Cyprus Institute of Neurology and Genetics. An experienced physiotherapist will perform all clinical assessment with exact the same methodological procedures.

#### 1. Mini Balance Evaluation Systems Test

It measures dynamic balance, functional mobility, and gait in neurological patients, including people with RRMS (80). The specific test consists of 14 items, including four of the six segments (anticipatory postural adjustments, sensory orientation, reactive postural control and dynamic gait) from the Balance Evaluation Systems Test. The Mini Balance Evaluation Systems Test should be scored out of 28 points to include 14 items that are scored from zero to two.

#### 2. Six Spot Step Test

It is a timed walking test that involves kicking over a number of targets placed along a 5m-path in which rely to some extent on vision and cognition (81). The Six Spot Step Test is measured in the time domain replicating a complex range of sensorimotor functions, part of which are lower limb strength, spasticity, coordination, as well as balance. We will perform the specific test as it is described by Nieuwenhuis et al (2006) (81) and we will record the mean time of the four runs as the final test result (82).

#### 3. Action Research Arm Test

It is a 19-item observational measure used by physiotherapists and other health care professionals to examine upper limb performance (i.e., coordination, dexterity and functioning) (83). Items covering the Action Research Arm Test are categorized into four subscales (grasp, grip, pinch and gross movement) and arranged in order of decreasing difficulty, with the most difficult task examined first, followed by the least difficult task. The patient is sitting comfortable in front of a stable desk performing each task and the performance is rated on a four-point scale, ranging from 0 (no movement) to 3 (movement performed normally). We will record the total score for each upper limb separately as the final test result.

#### 4. Isometric Dynamometer

We well assess the isometric muscle force of major muscle groups with the use of the muscle controller (Kinvent Biomechanique, Montpelier, France) which is a dynamometer used in the evaluation and rehabilitation of muscle strength that provides real time biofeedback (84). The patient lies (supine or prone) on a therapeutic bed and the physiotherapist, with the use of the muscle controller, holds against the patient’s limb as the patient exerts a maximal force. The physiotherapist counters the force (make test) or tries to break the contraction (break test) and the data will be stored using the KFORCE APP (Kinvent Biomechanique, Montpelier, France). Shoulder flexors, extensors, rotators, horizontal adductors and abductors adductors and abductors, elbow flexors and extensors are the major muscle groups which will be evaluated. A separate value for each muscle group will be recorded in order to be used in the visual and statistical analysis.

#### 5. Symbol Digit Modalities Test

We will employ the oral form which assesses the information processing speed (85). During the test, the participant will be given two minutes to orally match symbols with digits as quickly as possible. The key (specifying which symbols are assigned to which numbers) will be located at the top of a computer screen. The researcher will instruct the participants that each symbol is paired with a digit. Next, the participant will be instructed to perform the test by responding orally to each symbol. For example, the symbol “O” is matched with the number 6, so the correct response would be to say “six”. The researcher responsible for clinical assessments will record the participant’s responses directly on a computer screen. The score is obtained by subtracting the number of errors from the number of items completed in two minutes.

#### 6. Modified Fatigue Impact Scale

It is a short questionnaire which requires the participants to describe the effects of fatigue during the past four weeks (55) (S1. Appendix 1). The Modified Fatigue Impact Scale consists of 21 questions which are subjectively rated from “0” (low rate) to “4” (high rate) and it is also divided into three subscales (i.e., physical, cognitive, and psychosocial). We will record the total score of the test as the final test result. The higher the score is, the greater is the impact of fatigue in individual daily life. Although, to describe fatigue level the Fatigue Severity Scale is more commonly used, we decided not use it because it describes severe fatigue. Therefore, we will use the Modified Fatigue Impact Scale as the description of participants’ attribution of functional restrictions to fatigue symptoms.

### Analysis plan

To investigate possible effects of our protocol we will follow recommended guidelines (54), in which we will perform a separate analysis for each of the outcome measures, in all experimental phases (i.e., baseline, intervention and follow up). We will perform a visual analysis first, in order to determine whether there is a functional relationship between the intervention and the outcome measures, and secondly, we will perform a quantitative analysis methods to evaluate the magnitude of the intervention effect, provided there is evidence from the visual analysis (54). We will perform all neurophysiological and clinical assessments to each participant according to the number of data points during each phase (i.e., baseline, intervention, follow up) (Fig 1).

### TMS measures analysis

Corticospinal plasticity will be determined through changes of the corticospinal excitability and the central motor conduction time. Hence, we will quantify bilateral resting motor threshold, MEP amplitude and latency, because each measure can assess different plastic changes across the neuromotor axis and they can be used as a proxy of corticospinal plasticity. Resting motor threshold (% maximum stimulator output) states the general excitability of the neuromotor axis in the target muscle, amplitude (mV) expresses transynaptic activation of corticospinal neurons (86), while latency (ms) is the time between the TMS onset and the MEP onset, which refers to the integrity of the white matter fibers (87).

For both upper limbs, all neurophysiological measures (i.e., resting motor threshold, MEP amplitude, latency) will be first calculated from each MEP trace and then averaged to get a single value. These calculations will be done according to the different time points for each participant in the baseline phase, at five time points in the intervention phase and at three time points in the follow-up phase (Fig 1). In order to investigate possible changes in cortical excitability, we will measure resting motor threshold and peak-to-peak amplitude throughout assessing MEPs (88) of the Abductor Pollicis Brevis muscle, while measuring of latency will indicate possible changes in central motor conduction time. Any changes in all measures across time points, will indicate alterations in corticospinal plasticity (89). We will evaluate resting motor threshold using MTAT 2.0 (76) (available at http://clinicalresearcher.org/software.htm) and to investigate possible changes in individual corticospinal plasticity of each participant, we will calculate bilaterally the difference between the mean values of each phase (73,90). On the other hand, from each stimulus response during the suprathreshold stimulation (i.e., 120% of resting motor threshold) (89), we will calculate offline the MEP peak-to-peak amplitude and latency. To define central motor conduction time (ms), we will subtract the peripheral conduction time ((F wave latency + M wave latency – 1)/2) from the central conduction time (MEP latency). The F wave is the muscle response elicited by activation of the alpha motor neuron, while the M wave is the direct response of the muscle (66). A prolonged central motor conduction time indicates damage of large fibres, demyelination of central motor pathways or slow summation of descending excitatory potentials in the corticospinal tract evoked by TMS (66,69). To standardise all latencies (i.e., MEP, F and M wave), we will use a visual inspection from stimulation onset to response onset, performed from the same investigator so to ensure reliability of the study across all time points. To define possible changes in central motor conduction time, we will evaluate bilaterally the difference between the mean values of each phase.

### Clinical measures analysis

For each clinical measure (i.e., balance, gait, cognitive function, bilateral hand dexterity and strength) and for the Modified Impact Scale we will calculate the values from each time point across all phases (i.e., baseline, intervention, follow-up) and then we will evaluate the average of them, so to get a single mean value for each measure and for each phase (mean baseline; mean intervention mean follow-up). To investigate the association between the intervention protocol and clinical condition, we will calculate the differences between phases’ mean values (i.e., mean baseline; mean intervention; mean follow up), reflecting to the degree of change in clinical condition following in-phase bilateral exercises.

### Visual analysis

Two assessors will systematically measure each outcome measure across time, inter-assessor agreement will be calculated on at least twenty percent of the data points in each condition. The minimum acceptable inter-assessor agreement will be set to 0.8 (56).

Initially, a visual analysis will be conducted and presented graphically in a spaghetti plot, in order to define whether there is a functional relation between the intervention and the outcome measures (54). During the visual analysis, six features of the research design graphed data will be examined: level, trend, stability, immediacy of the effect, overlap, and consistency. Over the within-phase examination an evaluation of level, trend and stability will be examined. Level will be reported from the mean score of each dependent variable and trend will determine whether the data points are monotonically decreased or increased. Stability will be estimated based on the percentage of data points falling within 15% of the phase median, if this is higher than 80% then we assume that this criterion is met. Additionally, over the between-phase examination an evaluation of overlapping data among baseline and intervention phases, consistency of data patterns and immediacy of effect will be performed (54). The Percentage of Non-overlapping Data index will be used to quantify the proportion of data points in the intervention phase that do not overlap with the baseline phase (91) and the test statistic will be calculated using the Improvement Rate Difference as an effect size index. Immediacy of the effect will be examined by comparing changes in level between the last three data points of one phase and the three first data points of the next phase. Furthermore, consistency of data patterns involves the observation of the data from all phases within the same condition, with greater consistency expressing greater causal relation. Each feature will be assessed individually and collectively across to all participants and to all phases. Consequently, if the intervention protocol is the sole determinant of improvement, we expect to find indicators of improvement only at the intervention phase.

### Statistical analysis

For each of the outcome variables that the visual analysis indicates a potential functional effect, a quantitative analysis will be performed to estimate the effect size. This analysis will be performed for each variable at which a significant trend is shown as described in the previous section. In order to estimate the individual-level effect sizes, we will use three different methods, as suggested by ‘What Works Clearinghouse’ (56), the standardized mean difference (Cohen’s d), the standardized mean difference with correction for small sample sizes (Hedges’ g) and piecewise regression analysis which does not only reflect the immediate intervention effect, but also the intervention effect across time. Multilevel modelling, which is recommended by the ‘What Works Clearinghouse’ and the single case educational design, specific mean difference index will be used to estimate the magnitude of the effect across cases and compared to the effect obtained by the single level estimates (54).

For all the different neurophysiological parameters and clinical conditions, evaluated separately, the null hypothesis is that “there is no improvement from the proposed intervention”, thus participants’ responses are independent from the condition (baseline versus intervention) under which they were observed. The alternative hypothesis is that “the neurophysiological parameters and /or the clinical condition of the participants will be affected by the specific intervention”, again assessed separately. We will reject the null hypothesis if the p value is smaller than 0.05. All tests will be two sided. Statistical analysis will be performed using the statistical software R (https://www.r-project.org/).

## Data Availability

All relevant data from this study will be made available upon study completion.

## Supporting Information

**S1. Appendix 1. Modified Fatigue Impact Scale.**

**S2. Appendix 2. SPIRIT checklist**

**S3. Appendix 3. Study protocol**

## References

1. Lublin FD, Reingold SC. Defining the clinical course of multiple sclerosis: Results of an international survey. Neurology. 1996;46(4):907–11.

2. Moghaddam VK, Dickerson AS, Bazrafshan E, Seyedhasani SN. Socioeconomic determinants of global distribution of multiple sclerosis : an ecological investigation based on Global Burden of Disease data. 2021;1–11.

3. Walton C, King R, Rechtman L, Kaye W, Leray E, Marrie RA, et al. Rising prevalence of multiple sclerosis worldwide : Insights from the Atlas of MS, third edition. :1–6.

4. Lunde HMB, Assmus J, Myhr KM Bø L, Grytten N. Survival and cause of death in multiple sclerosis: A 60-year longitudinal population study. J Neurol Neurosurg Psychiatry. 2017;88(8):621–5.

5. Scalfari A, Knappertz V, Cutter G, Goodin DS, Ashton R, Ebers GC. Mortality in patients with multiple sclerosis. Neurology. 2013;81(2):184–92.

6. Kingwell E, Zhu F, Evans C, Duggan T, Oger J, Tremlett H. Causes that Contribute to the Excess Mortality Risk in Multiple Sclerosis: A Population-Based Study. Neuroepidemiology. 2020;54(2):131–9.

7. Dobson R, Giovannoni G. Multiple sclerosis –a review. Eur J Neurol. 2019;26(1):27–40.

8. Paz-Zulueta M, Parás-Bravo P, Cantarero-Prieto D, Blázquez-Fernández C, Oterino-Durán A. A literature review of cost-of-illness studies on the economic burden of multiple sclerosis. Mult Scler Relat Disord [Internet]. 2020;43(April):102162. Available from: https://doi.org/10.1016/j.msard.2020.102162

9. Maguire R, Maguire P. Caregiver Burden in Multiple Sclerosis: Recent Trends and Future Directions. Curr Neurol Neurosci Rep. 2020;20(7).

10. Kouzoupis AB, Paparrigopoulos T, Soldatos M, Papadimitriou GN. The family of the multiple sclerosis patient: A psychosocial perspective. Int Rev Psychiatry. 2010;22(1):83–9.

11. Lublin FD, Reingold SC, Cohen JA, Cutter GR, Sørensen PS, Thompson AJ, et al. Defining the clinical course of multiple sclerosis: The 2013 revisions. Neurology. 2014;83(3):278–86.

12. Lublin FD, Coetzee T, Cohen JA, Marrie RA, Thompson AJ. The 2013 clinical course descriptors for multiple sclerosis: A clarification. Neurology. 2020;94(24):1088–92.

13. Kister I, Bacon TE, Chamot E, Salter AR, Cutter GR, Kalina JT, et al. Multiple Sclerosis Symptoms. 2013;(June 2011):146–57.

14. Norbye AD, Midgard R, Thrane G. Spasticity, gait, and balance in patients with multiple sclerosis: A cross-sectional study. Physiother Res Int. 2020;25(1):1–9.

15. Benedict RHB, Amato MP, Deluca J, Geurts JJG. Cognitive impairment in multiple sclerosis : clinical management, MRI, and therapeutic avenues. Lancet Neurol [Internet]. 2020;19(10):860–71. Available from: http://dx.doi.org/10.1016/S1474-4422(20)30277-5

16. Frndak SE, Kordovski VM, Cookfair D, Rodgers JD, Weinstock-Guttman B, Benedict RHB. Disclosure of disease status among employed multiple sclerosis patients: Association with negative work events and accommodations. Mult Scler J. 2015;21(2):225–34.

17. Strober L, Chiaravalloti N, Moore N, Deluca J. Unemployment in multiple sclerosis (MS): Utility of the MS Functional Composite and cognitive testing. Mult Scler. 2014;20(1):112–5.

18. Fortune J, Norris M, Stennett A, Kilbride C, Lavelle G, Hendrie W, et al. Patterns and correlates of sedentary behaviour among people with multiple sclerosis: a cross-sectional study. Sci Rep [Internet]. 2021;11(1):1–10. Available from: https://doi.org/10.1038/s41598-021-99631-z

19. Lipp I, Tomassini V. Neuroplasticity and motor rehabilitation in multiple sclerosis. 2015;6(March):1–3.

20. Flachenecker P. Clinical implications of neuroplasticity - the role of rehabilitation in multiple sclerosis. Front Neurol. 2015;6(MAR):1–4.

21. Tomassini V, Matthews PM, Thompson AJ, Fuglø D, Geurts JJ, Johansen-berg H, et al. Neuroplasticity and functional recovery in multiple sclerosis. Nat Publ Gr [Internet]. 2012;8(11):635–46. Available from: http://dx.doi.org/10.1038/nrneurol.2012.179

22. Tavazzi E, Cazzoli M, Pirastru A, Blasi V, Rovaris M, Bergsland N, et al. Neuroplasticity and Motor Rehabilitation in Multiple Sclerosis : A Systematic Review on MRI Markers of Functional and Structural Changes. 2021;15(October).

23. Pawlitzki M, Neumann J, Heidel J, Stadler E, Sweeney-reed C, Sailer M. Loss of corticospinal tract integrity in early MS disease stages. 2017;0.

24. Fritz NE, Keller J, Calabresi PA, Zackowski KM. NeuroImage : Clinical Quantitative measures of walking and strength provide insight into brain corticospinal tract pathology in multiple sclerosis. NeuroImage Clin [Internet]. 2017;14:490–8. Available from: http://dx.doi.org/10.1016/j.nicl.2017.02.006

25. Bergsland N, Laganà MM, Tavazzi E, Caffini M, Tortorella P, Baglio F, et al. Corticospinal tract integrity is related to primary motor cortex thinning in relapsing-remitting multiple sclerosis. Mult Scler. 2015;21(14):1771–80.

26. Lemon RN. Descending pathways in motor control. Annu Rev Neurosci. 2008;31(Cm):195–218.

27. Warraich Z, Kleim JA. Neural plasticity: The biological substrate for neurorehabilitation. PM R [Internet]. 2010;2(12 SUPPL):S208–19. Available from: http://dx.doi.org/10.1016/j.pmrj.2010.10.016

28. Mori F, Kusayanagi H, Nicoletti CG, Weiss S, Marciani MG, Centonze D. Cortical plasticity predicts recovery from relapse in multiple sclerosis. Mult Scler J. 2014;20(4):451–7.

29. Mori F, Rossi S, Piccinin S, Motta C, Mango D, Kusayanagi H, et al. Synaptic plasticity and PDGF signaling defects underlie clinical progression in multiple sclerosis. J Neurosci. 2013;33(49):19112–9.

30. Pascual-Leone a, Tarazona F, Keenan J, Tormos JM, Hamilton R, Catala MD. Transcranial magnetic stimulation and neuroplasticity. Neuropsychologia [Internet]. 1999;37(2):207–17. Available from: http://www.ncbi.nlm.nih.gov/pubmed/10080378

31. Neva JL, Lakhani B, Brown KE, Wadden KP, Mang CS, Ledwell NHM, et al. Multiple measures of corticospinal excitability are associated with clinical features of multiple sclerosis. Behav Brain Res [Internet]. 2016;297:187–95. Available from: http://dx.doi.org/10.1016/j.bbr.2015.10.015

32. Bestmann S, Krakauer JW. The uses and interpretations of the motor - evoked potential for understanding behaviour. 2015;679–89.

33. Udupa K, Chen R. Central motor conduction time [Internet]. 1st ed. Vol. 116, Handbook of Exploration and Environmental Geochemistry. Elsevier B.V.; 2013. 375–386 p. Available from: http://dx.doi.org/10.1016/B978-0-444-53497-2.00031-0

34. Zeller D, Classen J. Plasticity of the motor system in multiple sclerosis. Neuroscience [Internet]. 2014;283(June):222–30. Available from: http://dx.doi.org/10.1016/j.neuroscience.2014.05.043

35. Zentgraf K, Helm F. Brain Changes in Response to Exercise - Methodologies for Identifying the Physiological Effects of Physical Exercise. 2020;II:815–31.

36. Prosperini L, Filippo M Di. Beyond clinical changes: Rehabilitation-induced neuroplasticity in MS. Mult Scler J. 2019;25(10):1348–62.

37. Moucha R Ã MPK. Cortical plasticity and rehabilitation. 2006;

38. Diechmann MD, Campbell E, Coulter E, Paul L, Dalgas U, Hvid LG. Effects of exercise training on neurotrophic factors and subsequent neuroprotection in persons with multiple sclerosis—a systematic review and meta-analysis. Brain Sci. 2021;11(11).

39. Sandroff BM, Jones CD, Baird JF, Motl RW. Systematic Review on Exercise Training as a Neuroplasticity-Inducing Behavior in Multiple Sclerosis. Neurorehabil Neural Repair. 2020;34(7):575–88.

40. Learmonth YC, Motl RW. Exercise Training for Multiple Sclerosis : A Narrative Review of History, Benefits, Safety, Guidelines, and Promotion. 2021;

41. Sun Y, Zehr EP. Training-induced neural plasticity and strength are amplified after stroke. Exerc Sport Sci Rev. 2019;47(4):223–9.

42. Garry MI, van Steenis RE, Summers JJ. Interlimb coordination following stroke. Hum Mov Sci. 2005;24(5–6):849–64.

43. Whitall J, McCombe Waller S, Sorkin JD, Forrester LW, Macko RF, Hanley DF, et al. Bilateral and unilateral arm training improve motor function through differing neuroplastic mechanisms: A single-blinded randomized controlled trial. Neurorehabil Neural Repair. 2011;25(2):118–29.

44. Smith AL, Richard Staines W. Cortical and behavioral adaptations in response to short-term inphase versus antiphase bimanual movement training. Exp Brain Res. 2010;205(4):465–77.

45. Neva JL, Legon W, Staines WR. Primary motor cortex excitability is modulated with bimanual training. Neurosci Lett [Internet]. 2012;514(2):147–51. Available from: http://dx.doi.org/10.1016/j.neulet.2012.02.075

46. Stinear JW, Byblow WD. Disinhibition in the human motor cortex is enhanced by synchronous upper limb movements. J Physiol. 2002;543(1):307–16.

47. Liepert J, Mingers D, Heesen C, Bäumer T, Weiller C. Motor cortex excitability and fatigue in multiple sclerosis: A transcranial magnetic stimulation study. Mult Scler. 2005;11(3):316–21.

48. Toyokura M, Muro I, Komiya T, Obara M. Activation of pre-supplementary motor area (SMA) and SMA proper during unimanual and bimanual complex sequences: An analysis using functional magnetic resonance imaging. J Neuroimaging. 2002;12(2):172–8.

49. Staines WR, McIlroy WE, Graham SJ, Black SE. Bilateral movement enhances ipsilesional cortical activity in acute stroke: A pilot functional MRI study [4] (multiple letters). Neurology. 2001;57(9):1740–1.

50. Reina-Gutiérrez S, Cavero-Redondo I, Martínez-Vizcaíno V, Núñez de Arenas-Arroyo S, López-Muñoz P, Álvarez-Bueno C, et al. The type of exercise most beneficial for quality of life in people with multiple sclerosis: A network meta-analysis. Ann Phys Rehabil Med. 2022;65(3).

51. Akbar N, Sandroff BM, Wylie GR, Strober LB, Smith A, Goverover Y, et al. Progressive resistance exercise training and changes in resting-state functional connectivity of the caudate in persons with multiple sclerosis and severe fatigue: A proof-of-concept study. Neuropsychol Rehabil [Internet]. 2020;30(1):54–66. Available from: https://doi.org/10.1080/09602011.2018.1449758

52. Proschinger S, Kuhwand P, Rademacher A, Walzik D, Warnke C, Zimmer P, et al. Fitness, physical activity, and exercise in multiple sclerosis : a systematic review on current evidence for interactions with disease activity and progression. J Neurol [Internet]. 2022;(January). Available from: https://doi.org/10.1007/s00415-021-10935-6

53. Calabrese M, Filippi M, Gallo P. Cortical lesions in multiple sclerosis. Nat Rev Neurol [Internet]. 2010;6(8):438–44. Available from: http://dx.doi.org/10.1038/nrneurol.2010.93

54. Lobo MA, Moeyaert M, Cunha AB, Babik I. Single-case design, analysis, and quality assessment for intervention research. J Neurol Phys Ther. 2017;41(3):187–97.

55. Fisk JD, Ritvo PG, Ross L, Haase DA, Marrie TJ, Schlech WF. Measuring the functional impact of fatigue: Initial validation of the fatigue impact scale. Clin Infect Dis. 1994;18:S79–83.

56. Kratochwill, T. R. Hitchcock, J. Horner, R. H. Levin, J. R. Odom, S. L. Rindskopf, D. M Shadish WR. Single‐Case Design Technical Documentation. Work Clear website http://ies.ed.gov/ncee/wwc/pdf/wwc_scd.pdf. 2010;(mDecember):2010.

57. Zhan S, Ottenbacher KJ. Single subject research designs for disability research. Disabil Rehabil. 2001;23(1):1–8.

58. Kurtzke JF, Kurtzke JF. Rating neurologic impairment in multiple sclerosis : An expanded disability status scale (EDSS). 1983;

59. Folstein MF, Folstein SE, McHugh PR. “Mini-mental state”. A practical method for grading the cognitive state of patients for the clinician. J Psychiatr Res. 1975;12(3):189–98.

60. Meseguer-Henarejos AB, Sânchez-Meca J, López-Pina JA, Carles-HernâNdez R. Inter-and intra-rater reliability of the Modified Ashworth Scale: A systematic review and meta-analysis. Eur J Phys Rehabil Med. 2018;54(4):576–90.

61. Kalb R, Brown TR, Coote S, Costello K, Dalgas U, Garmon E, et al. Exercise and lifestyle physical activity recommendations for people with multiple sclerosis throughout the disease course. Mult Scler J. 2020;26(12):1459–69.

62. Adler SS, Beckers D, Buck M. PNF in Practice. PNF Pract. 2000;

63. Abbaspoor E, Zolfaghari M, Ahmadi B, Khodaei K. The effect of combined functional training on BDNF, IGF-1, and their association with health-related fitness in the multiple sclerosis women. Growth Horm IGF Res [Internet]. 2020;52(March):101320. Available from: https://doi.org/10.1016/j.ghir.2020.101320

64. Hermens HJ, Freriks B, Hermens HJ, Freriks B. Future Applications of Surface ElectroMyoGraphy. 1999;(September):4–5.

65. Hidasi E, Diószeghy P, Csépány T, Mechler F, Bereczki D. Peripheral nerves are progressively involved in multiple sclerosis - A hypothesis from a pilot study of temperature sensitized electroneurographic screening. Med Hypotheses [Internet]. 2009;72(5):562–6. Available from: http://dx.doi.org/10.1016/j.mehy.2008.07.066

66. Hallett M. Transcranial Magnetic Stimulation: A Primer. Neuron. 2007;55(2):187–99.

67. McNeil CJ, Butler JE, Taylor JL, Gandevia SC. Testing the excitability of human motoneurones. Front Hum Neurosci. 2013;7(APR 2013):1–9.

68. Fisher MA. F-waves - Physiology and clinical uses. ScientificWorldJournal. 2007;7:144–60.

69. Zimnowodzki S, Butrum M, Kimura J, Stålberg E, Mahajan S, Gao L. Emergence of F-waves after repetitive nerve stimulation. Clin Neurophysiol Pract [Internet]. 2020;5:100–3. Available from: https://doi.org/10.1016/j.cnp.2020.04.002

70. Groppa S, Oliviero A, Eisen A, Quartarone A, Cohen LG, Mall V, et al. A practical guide to diagnostic transcranial magnetic stimulation: Report of an IFCN committee. Clin Neurophysiol [Internet]. 2012;123(5):858–82. Available from: http://dx.doi.org/10.1016/j.clinph.2012.01.010

71. Rossini PM, Burke D, Chen R, Cohen LG, Daskalakis Z, Di Iorio R, et al. Non-invasive electrical and magnetic stimulation of the brain, spinal cord, roots and peripheral nerves: Basic principles and procedures for routine clinical and research application: An updated report from an I.F.C.N. Committee. Clin Neurophysiol [Internet]. 2015;126(6):1071–107. Available from: http://dx.doi.org/10.1016/j.clinph.2015.02.001

72. Pascual-Leone A, Dang N, Cohen LG, Brasil-Neto JP, Cammarota A, Hallett M. Modulation of muscle responses evoked by transcranial magnetic stimulation during the acquisition of new fine motor skills. J Neurophysiol. 1995;74(3):1037–45.

73. Balloff C, Penner I-K, Ma M, Georgiades I, Scala L, Troullinakis N, et al. The degree of cortical plasticity correlates with cognitive performance in patients with Multiple Sclerosis. Brain Stimul [Internet]. 2022;15(2):403–13. Available from: https://doi.org/10.1016/j.brs.2022.02.007

74. Charalambous CC, Dean JC, Adkins DAL, Hanlon CA, Bowden MG. Characterizing the corticomotor connectivity of the bilateral ankle muscles during rest and isometric contraction in healthy adults. J Electromyogr Kinesiol [Internet]. 2018;41(February):9–18. Available from: https://doi.org/10.1016/j.jelekin.2018.04.009

75. Rossini PM, Barker AT, Berardelli A, Caramia MD, Caruso G, Cracco RQ, et al. Non-invasive electrical and magnetic stimulation of the brain, spinal cord, roots and peripheral nerves: Basic principles and procedures for routine clinical and research application: An updated report from an I.F.C.N. Committee. Clin Neurophysiol. 1994;91(2):79–92.

76. Awiszus F. TMS and threshold hunting [Internet]. Vol. 56. Elsevier B.V.; 2003. 13–23 p. Available from: http://dx.doi.org/10.1016/S1567-424X(09)70205-3

77. Silbert BI, Patterson HI, Pevcic DD, Windnagel KA, Thickbroom GW. Clinical Neurophysiology A comparison of relative-frequency and threshold-hunting methods to determine stimulus intensity in transcranial magnetic stimulation. Clin Neurophysiol [Internet]. 2013;124(4):708–12. Available from: http://dx.doi.org/10.1016/j.clinph.2012.09.018

78. Goldsworthy MR, Hordacre B, Ridding MC. Minimum number of trials required for within- and between-session reliability of TMS measures of corticospinal excitability. Neuroscience [Internet]. 2016;320:205–9. Available from: http://dx.doi.org/10.1016/j.neuroscience.2016.02.012

79. Snow NJ, Wadden KP, Chaves AR, Ploughman M. Review Article Transcranial Magnetic Stimulation as a Potential Biomarker in Multiple Sclerosis : A Systematic Review with Recommendations for Future Research. 2019;2019.

80. Franchignoni F, Horak F, Godi M, Nardone A, Giordano A. Using psychometric techniques to improve the balance evaluation systems test: The mini-bestest. J Rehabil Med. 2010;42(4):323–31.

81. Nieuwenhuis MM, Tongeren H Van, Sørensen PS, Ravnborg M. The Six Spot Step Test : a new measurement for walking ability in multiple sclerosis. 2006;(September 2005).

82. Callesen J, Richter C, Kristensen C, Sunesen I, Næsby M, Dalgas U, et al. Test–retest agreement and reliability of the Six Spot Step Test in persons with multiple sclerosis. Mult Scler J. 2019;25(2):286–94.

83. Carpinella I, Cattaneo D, Ferrarin M. Quantitative assessment of upper limb motor function in Multiple Sclerosis using an instrumented Action Research Arm Test. J Neuroeng Rehabil. 2014;11(1):1–16.

84. Andrews AW, Thomas MW, Bohannon RW. Normative values for isometric muscle force measurements obtained with hand-held dynamometers. Phys Ther. 1996;76(3):248–59.

85. Benedict RHB, Deluca J, Phillips G, LaRocca N, Hudson LD, Rudick R. Validity of the Symbol Digit Modalities Test as a cognition performance outcome measure for multiple sclerosis. Mult Scler. 2017;23(5):721–33.

86. Ziemann U, Reis J, Schwenkreis P, Rosanova M, Strafella A, Badawy R, et al. TMS and drugs revisited 2014. Clin Neurophysiol [Internet]. 2015;126(10):1847–68. Available from: http://dx.doi.org/10.1016/j.clinph.2014.08.028

87. Hess CW, Mills KR, Murray NM. Responses in small hand muscles from magnetic stimulation of the human brain. J Physiol. 1987;388(1):397–419.

88. Vanteemar S. Sreeraj S, Uvais2 NA, Mohanty3 S, Kumar3 S, Department. Indian nursing students’ attitudes toward mental illness and persons with mental illness. Ind Psychiatry J. 2019;195–201.

89. Paulus W, Peterchev A V., Ridding M. Transcranial electric and magnetic stimulation: Technique and paradigms. Handb Clin Neurol. 2013;116(0):329–42.

90. Stampanoni Bassi M, Buttari F, Maffei P, De Paolis N, Sancesario A, Gilio L, et al. Practice-dependent motor cortex plasticity is reduced in non-disabled multiple sclerosis patients. Clin Neurophysiol [Internet]. 2020;131(2):566–73. Available from: https://doi.org/10.1016/j.clinph.2019.10.023

91. Heyvaert M, Onghena P. Analysis of single-case data: Randomisation tests for measures of effect size. Neuropsychol Rehabil. 2014;24(3–4):507–27.

